# Transition from Acute to Chronic Low Back Pain in a Community-Based Cohort

**DOI:** 10.1101/2024.03.19.24304350

**Authors:** Colleen Burke, Rebecca Fillipo, Steven Z. George, Adam P. Goode

## Abstract

The transition from acute to chronic low back pain (LBP) in community settings has yet to be well understood. We recruited n=131 participants with acute LBP from the community and followed them for 3 and 6-month outcomes. Acute LBP was defined by a duration of < 4 weeks, and participants must have reported a 30-day LBP-free period before the current acute episode. Chronic LBP was defined as pain most or every day over the past 3 months. Baseline psychological, social, and demographic factors were examined as predictors of transition to chronic LBP at 3 months or continuation of chronic LBP at 6 months. The transition from acute to chronic LBP at 3 months was 32.2% ( 38/118). At 6 months, 80.7% (25/111) of participants who transitioned at 3 months continued to have chronic LBP. At 6 months, participants who identified as Black or African American were more likely than white participants to transition to chronic LBP (RR=1.76, 95% CI 1.05, 2.95) and more likely to continue to have chronic LBP (RR=2.19, 95% CI 1.14, 4.21). Those classified at baseline by both LBP most or every day and intensity of at least 30/100 were more likely to transition to chronic LBP (RR=3.13, 95% CI 1.84, 5.30) and continue to have chronic LBP at 6 months (RR=2.58, 95% CI 1.43, 4.16). The STarT Back Screening Tool and the OSPRO-YF were associated with the transition to chronic LBP at 3 months and continuation at 6 months. Participants with higher PROMIS General Health and PROMIS Physical Health scores were less likely to transition to chronic LBP or continue to have chronic LBP at 6 months. These findings identify factors of acute LBP in the community that may predict the transition to chronic LBP. Larger studies are needed to confirm these findings and better understand the mechanisms driving the transition to chronic LBP.

## Introduction

Acute low back pain (LBP) is a common occurrence, with up to 25% of individuals each year reporting an event. [47; 51] and as many as 63% will transition to chronic LBP. [34; 47; 50; 51] Chronic LBP, characterized by pain most or every day for three months, [57] is the leading cause of years lived with disability [13; 31] and is a costly ($28.2 billion per year) medical problem in the United States [14]. [11; 12; 41] Despite the significance, the exact reasons individuals transition from acute to chronic LBP are mostly unknown but are thought to be a complex relationship between biopsychosocial factors. [53] Many psychological factors, such as depression, anxiety, and catastrophizing, as well as social factors, such as race, gender, socioeconomic position, and lifestyle, are associated with chronic LBP in care-seeking populations. [35] However, psychological characteristics and social risk factors involved in the transition to chronic LBP are relatively understudied and mainly absent from the community.

Previous work has investigated chronic, recurrent, and continuous pain[54; 55], but the classification of acute pain or pain before the chronic stage has been limited. Recently, Eccleston proposed a framework for studying acute pain and the transition to chronic pain.[18] Applying this framework in the community setting, especially in the context of psychosocial factors, may elucidate factors associated with the transition to chronic LBP. A notable limitation of the existing literature is that for studies estimating chronic LBP incidence, it is typical to use cohorts restricted to patients seeking healthcare. However, a large proportion (42%) of individuals experiencing LBP do not seek care.[22] Because of this, results from care-seeking cohorts may not be generalizable to adults with acute LBP. Cohorts recruited directly from the community (i.e., not limited to individuals seeking care) play an essential role in understanding the full spectrum of health and disease processes regardless of healthcare access or utilization. For example, a recent Australian community-based cohort study indicated that acute LBP’s prognosis is far better than in clinical populations.[15] However, the same may not be accurate in the US, given the differences between these countries (e.g., demographic makeup, sociopolitical, and healthcare characteristics). The limited US community-based studies also indicate that chronic LBP incidence is higher than previously thought (approximately 25%).[25; 51]

More studies enrolling participants directly from the community are needed to understand better the transition from acute to chronic LBP and the potential differences between sociodemographic and psychological measures in the transition from acute to chronic LBP. To fill these gaps, we conducted a preliminary study using a US-based cohort of participants recruited from the community with acute LBP. We followed them over time for 3 and 6 month follow-up measurements. The purpose of this manuscript is to characterize the trajectory of participants who have an acute LBP episode over 6 months and determine the baseline demographic, psychological, sociological, pain, and health characteristics associated with the transition to chronic LBP at 3 months and reporting continued chronic LBP symptoms at 6 months.

## Methods

We used data from an ongoing cohort study of community-based adults investigating the biopsychosocial factors related to transitioning from new-onset acute LBP to chronic LBP. Study participants were recruited from communities in and around Durham, NC and Kannapolis, NC between February 2022 and November 2022. We utilized a community-based approach to recruit potential participants at both sites; this included advertisements on social media, newspaper articles, volunteer registries, emails distributed through university networks, flyers posted in or around the communities, and word of mouth. Recruitment in Kannapolis was primarily based on the MURDOCK Study. [7] The MURDOCK Study is a 12,526-participant community-based longitudinal cohort recruited from 2007-2013 centralized in Cabarrus County. In Durham County, we recruited participants using social media, flyers at local events, and the Duke Health Volunteer Registry. This registry can be queried for inclusion and exclusion criteria related to age, comorbidities (cancer/autoimmune conditions), and recent surgery/trauma. It contains over 7,200 volunteers accessible by email, and among those 55% are healthy, 56% identify as White, 35% identify as Black, and 5% identify as Hispanic. This study was reviewed and approved by the institutional review board at the Duke University School of Medicine.

To be eligible for study participation, individuals were required to be adults (≥18 years old) with acute LBP: LBP that started <4 weeks prior to screening and ≥30 days without LBP before the date of acute onset. We excluded individuals with a current or previous history of systemic inflammatory or autoimmune conditions (i.e., rheumatoid arthritis, spondyloarthropathies, irritable bowel syndrome, multiple sclerosis, Guillain-Barré Syndrome, human immunodeficiency virus, lupus), cancer (other than skin cancer), lumbar spine surgery, low back trauma (e.g., motor vehicle accident, falls), and congenital or acquired spinal defect (e.g., scoliosis). We also excluded individuals who were pregnant. Eligible individuals received and signed an informed consent form electronically via Research Electronic Data Capture (REDCap)[27; 28; 42] or in person. All participants were provided a copy of their signed consent form and scheduled for a baseline in-person visit no later than 6 weeks from the date of LBP onset. Before in-person data collection, participants completed study questionnaires to assess pain intensity, interference, and duration; depression, anxiety, and social measures via an online questionnaire using REDCap or, if the participant preferred, by phone. Study staff reviewed procedures with participants at the start of the in-person data collection visit. Participants were compensated for each in-person data collection visit and completion of electronic questionnaires.

### Chronic Low Back Pain Outcome Definition

The primary outcome was chronic LBP at 3 months. We also examined the continuation of chronic LBP from 3 months to 6 months as an outcome. Chronic LBP was defined and categorized by the Graded Chronic Pain Scale-Revised[56] with the definition of chronic LBP at 3 and 6 months, LBP most or every day for the past 3 months.

Participants who indicated they had pain most or every day for the past three months were classified as having chronic LBP. This scale further categorizes chronic pain based on both frequencies of pain (i.e., never, somedays, most days, or every day) and interference with daily work or life activities to define high-impact chronic pain at 3 months. High-impact chronic LBP was further defined as pain that limited activities most or every day in the past 3 months. Bothersome and mild chronic pain are defined using the frequency of pain interference from the Pain, Enjoyment of Life and General Activities (PEG) scale[37]. Bothersome chronic LBP was defined as pain that interfered with work or life activities ‘some days’ or ‘never’ and a PEG score of ≥12, while mild chronic LBP was defined as pain that interfered with work or life activities ‘some days’ or ‘never’ and a PEG score of <12. We also measured potential recurrence of LBP at both the 3-month and 6-month time points by asking participants the following question “In the past 3-months, have you had a period of at least 30 days without low back pain?”.

### Sociodemographic Characteristics

We collected self-reported sociodemographic characteristics, including age, sex at birth, gender identity, racial identity, Hispanic ethnicity (yes/no), highest educational attainment, insurance type, marital status, and employment status. Participants could indicate one or more racial identities out of the following categories: American Indian or Alaskan Native, Asian, Black or African American, Native Hawaiian or Other Pacific Islander, or White. Participants also had the option to select “other” and provide a write-in racial category option or select “unknown” or “choose not to respond.” To assess the presence of racial disparities, we grouped participants into two groups: 1) racially minoritized[26], which included participants who identified as Asian, Black or African American, and those who had selected more than one race category (there were no participants who identified as American Indian or Alaskan Native, or Native Hawaiian or Other Pacific Islander), and 2) participants who racially identified as White.[8] Participants reported sex at birth (female/male) and gender identity (cisgender, transgender, non-binary, genderqueer, agender, or gender fluid) using standardized items.[3]

### Pain Characteristics

Beyond the questions used to categorize acute LBP (duration of LBP symptoms <4 weeks duration), we collected information regarding other pains and LBP treatments used since the onset of current LBP (adapted from the NIH recommended minimum dataset for chronic LBP),[16] personal history of prior LBP episodes, and family history of LBP and chronic pain. [30]We measured pressure pain threshold (PPT) at the upper trapezius (PPT-UT) and posterior superior iliac spine (PPT-PSIS) bilaterally using a standard rubber-tip algometer. All PPT tests were performed with participants seated with their hands in their lap. Study staff explained the procedure to the participants and instructed them to say “pain” as soon as the pressure applied first produced a painful sensation. Pressure was applied at a rate of 1-kgf/cm2/second until the participant’s PPT was reached up to 10.1 kgf. If the PPT was not reached before this ceiling, the measurement was recorded at a truncated value of 10.1 kgf. Three measurements for PPT-UT and PPT-PSIS were recorded on each side (alternating between left and right); we present the mean value (in kgf) of the PPT-UT and PPT-PSIS measurements.

### General Health, Clinical and Social Health Characteristics

We used the General Health Item from the Patient Reported Outcomes Measurement Information System (PROMIS) Scale v1.2 – Global Health [29] to capture self-reported health status: Excellent, Very Good, Good, Fair, or Poor.

Participant height and weight were collected in person on a standard electronic scale and used to calculate body mass index (BMI). We captured two components of social health: social isolation and the ability to perform social roles. We ascertained social isolation using the Social Network Index (SNI), [4] as recommended by the Institute of Medicine (IOM) [24; 43] The SNI asks questions about the frequency of contact with friends and family, religious participation, group membership, and marital status to classify individuals into four mutually exclusive groups: most isolated, very isolated, somewhat isolated, and not isolated.[24; 29] We used two items from the PROMIS scale v1.2—Global Health measure to specifically capture social function, one assessing the performance of social activities and a second item assessing the satisfaction with social roles.[29] Stress symptoms were collected using the validated single-item question: “Stress means a situation in which a person feels tense, restless, nervous, or anxious, or is unable to sleep at night because their mind is troubled all the time. Do you feel this kind of stress these days?” as recommended by the IOM[24] Participants are given the following response options: Not at All, A Little Bit, Somewhat, Quite a Bit, and Very Much.[19; 24]

### Psychological Characteristics

To capture depressive symptoms, we collected the PROMIS Short Form v1.0 – Depression 4a.[45; 46] [19; 24]We also collected the STarT Back Screening Tool (SBT)[30]. This 9-item questionnaire asks about LBP spreading down the leg(s), pain in other areas, perception of physical activity safety, pain catastrophizing, and LBP-related functional limitations to categorize individuals with LBP into low-, medium-, and high-risk categories based on their predicted risk of acute-to-chronic LBP transition. We used the Optimal Screening for Prediction of Referral and Outcome Yellow Flags (OSPRO-YF) 10-item questionnaire to measure composite psychological distress. The OSPRO-YF is a validated tool that estimates scores on the following measures: Fear-Avoidance Belief Questionnaire (both physical activity and work subscales; FABQ-PA and FABQ-W), Pain Anxiety Symptoms Scale (PASS-20), Pain Catastrophizing Scale (PCS), Patient Health Questionnaire-9 (PHQ-9), Pain Self-Efficacy Questionnaire (PSEQ), State-Trait Anxiety Inventory (STAI), State-Trait Anger Expression Inventory (STAXI), and Tampa Scale of Kinesiophobia (TSK-11).[10; 23; 39]

### Definitions of Acute Low Back Pain

Acute LBP for inclusion in the study was based on the duration of symptoms for the current episode. Based upon our prior work on acute LBP, we examined the frequency, intensity, and interference of pain at baseline. From these measures, we proposed three unique definitions of acute LBP: impact-based, intensity-based (with Visual Analogue Scale (VAS) cut-points at 20-, 30-, and 40/100), and interference-based definitions; each definition relied on participants’ responses to two items. The impact-based definitions used the questions: 1) “Since the onset of your pain, how often have you had pain? Would you say Never, Some Days, Most Days, or Every Day?” and 2) “Since the onset of your pain, what number best describes how pain has interfered with your general activity?”. The intensity-based definition combined LBP frequency “Since the onset of your pain, how often have you had pain? Would you say Never, Some Days, Most Days, or Every Day?” with LBP intensity using the 0-100 VAS (and cut-points at 20-, 30-, and 40-were examined). Finally, the interference-based definition used the following two questions: 1) “Since the onset of your pain, how often have you had pain? Would you say Never, Some Days, Most Days, or Every Day?” and 2) the following PEG item: “Since the onset of your pain, what number best describes how pain has interfered with your general activity?” Responses to these questions were used to classify participants’ acute LBP experience into one of three ranked categories.

### Statistical Analysis

We conducted all analyses in SAS 9.4 (Cary, NC) or Stata 18 (College Station, TX) and used RStudio for data visualization.[32; 48; 58] SankeyMATIC online software was used to develop the illustration of the transition from baseline to 3 months and 6 months.[9] Counts, proportions, means, and standard deviations (SD) were used for univariate descriptions. Student t-tests and chi-square tests were used to determine differences in continuous and categorical variables across both 3- and 6 month LBP outcomes. We used Fisher’s exact tests when cell counts for categorical variables were below n=5. Unadjusted risk ratios (RR) and 95% confidence intervals (CI) were estimated for each variable and 3- and 6-month outcomes. Some binomial models did not achieve convergence, in which case a logit link was used for estimation. Chronic LBP at 3 and 6 months for our models is based upon the respondents having LBP most or everyday in the past 3 months. We also describe the sample size and frequency of those participants that met the definition of high impact, bothersome, and mild chronic LBP at 3 and 6 months. Participants (n = 5) who transitioned to chronic LBP between 3 and 6 months did not meet our definitions for “transition” or “continuation” of LBP, so we removed those participants for bivariate comparisons to estimate the relationship between each variable and continued chronic LBP at 6 months.

## Results

We screened 384 potential participants, 184 of whom met the study criteria. The most common reason for exclusion was a duration of LBP >4 weeks or having LBP within 30 days before the current LBP episode (158/200 (79%)), followed by history of cancer (16/200 (8%)), active inflammatory condition (11/200 (5.5%)), history of lumbar surgery (7/200 (3.5%)) and other reasons (8/200 (4%)). Of the 184 people who screened eligible, 143 (77.7%) enrolled in the study, and of those, 131 (91.6%) enrolled provided baseline data. No significant differences in the distribution of age, racial identity, or sex at birth were found between those eligible and those who did not enroll and those who did enroll in the study or those who did and did not provide baseline data.

We present sociodemographic characteristics for the overall sample in **Table 1**. Sex at birth for much of our sample (58.8%) was female, and 100% of our sample had cisgender identity. Mean (SD) age was 56.9 (13.8), ranging from 19 to 81 years old. Regarding race, 22.9% of participants self-identified as Black or African American, and 67.9% identified as White. There were 6.1% of participants (of any race) who reported Hispanic ethnicity. The most common employment status was working full-time (42.7%), with 42.0% of all participants reporting having some college, and 51.2% having a bachelor’s degree or higher. A large proportion (72.5%) reported never smoking, whereas 7.6% were current smokers (smoking some or every day).

**Table 1.**
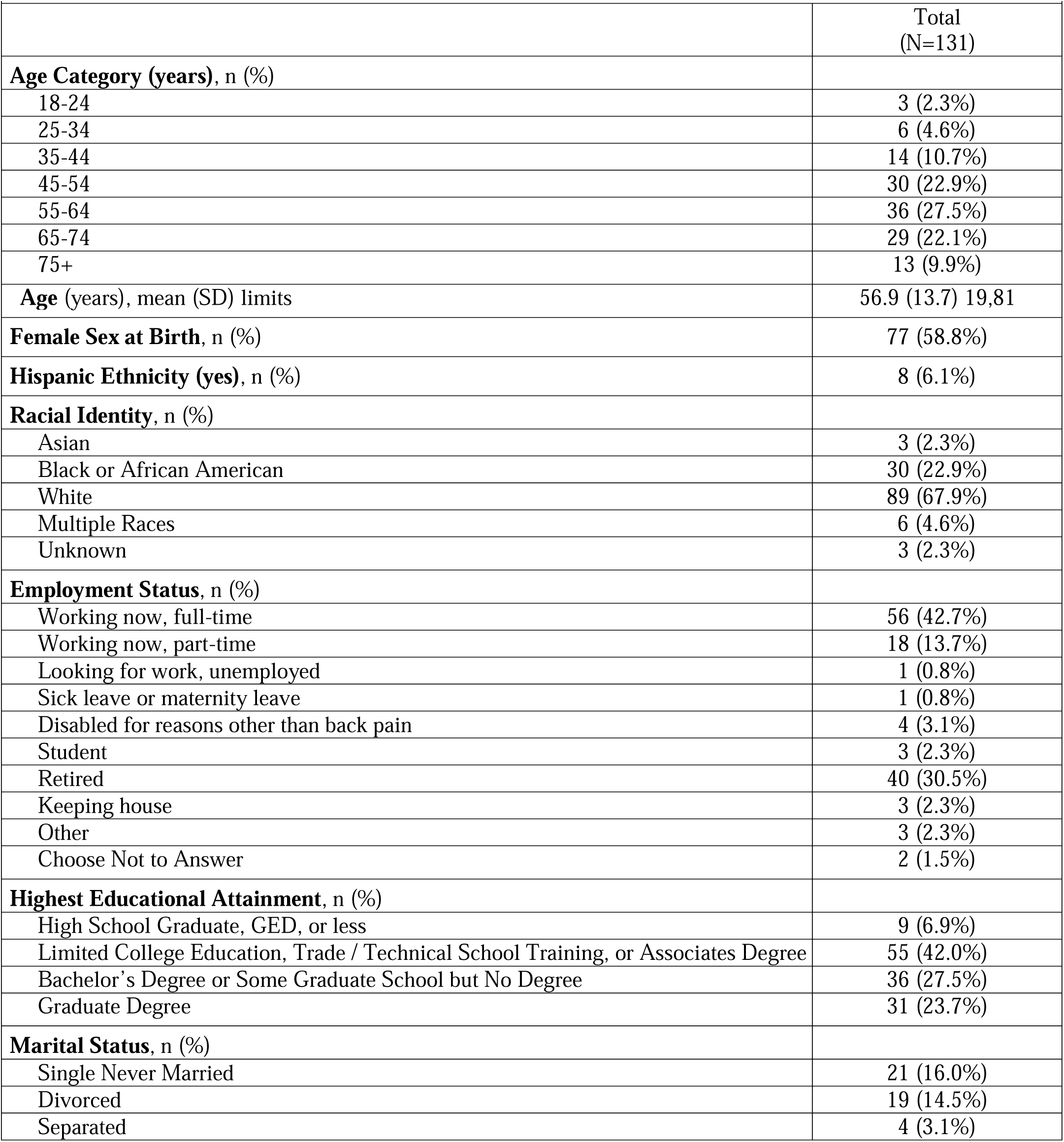
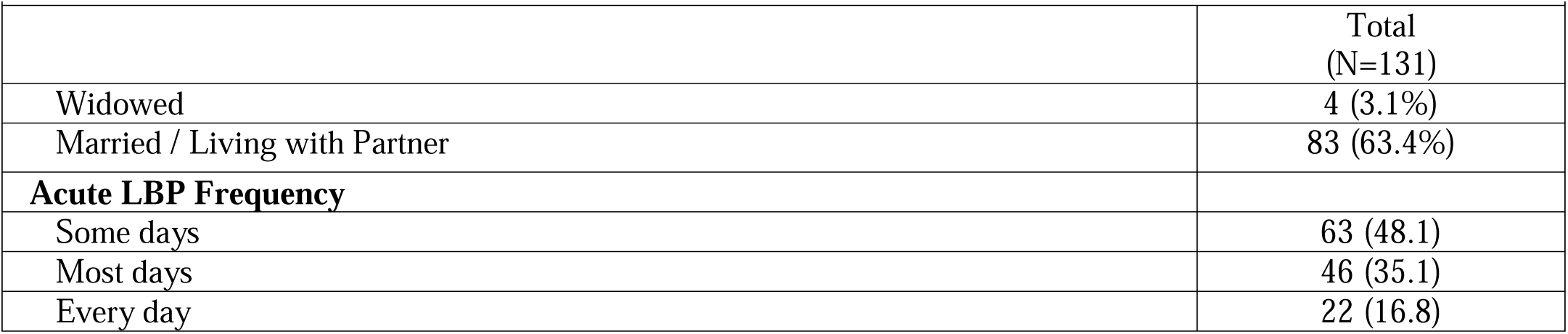
Characteristics of the Cohort at Baseline.

We present the transition to chronic LBP counts and proportions in **Table 2**. The proportion of participants who transitioned to chronic LBP, low back pain most or every day for the past 3 months, at 3 months was 32.2% (38/118).

**Table 2.** The transition from acute to chronic at 3 and 6 months.

|  | 3-months (n=118) | 6-months (n=111) |
| --- | --- | --- |
| Chronic LBP, n (%) | 38 (32.2%) | 31 (27.9%) |
| High-Impact, n (%) | 4 (10.5%) | 8 (25.8%) |
| Bothersome | 22 (57.9%) | 13 (41.9%) |
| Mild | 12 (31.6%) | 10 (32.3%) |
| Continued Chronic LBP | N/A | 25 (80.7%) |
| New Chronic | N/A | 6 (19.3%)* |
| LBP=low back pain. Chronic LBP defined as having LBP most or every day in the past 3 months. Chronic, high-impact, bothersome and mild chronic low back pain defined by the Graded Chronic Pain Scale Revised. Continued chronic LBP=those participants with chronic LBP at 3 months and continued to meet the definition at 6 months. New chronic LBP=new chronic LBP at 6 months. *N=5 participants transitioned to chronic LBP at 6 months and n=1 participant was missing 3 month outcomes but completed 6 months outcomes. |  |  |

Among those who transitioned, 10.5% (4/118) had high-impact chronic LBP, 52.6% (22/118) had bothersome chronic LBP, and 36.8% (12/118) had mild chronic LBP. At 6 months, a large majority (80.7%, 25/111) of participants continued to have chronic LBP, and the proportion of participants with high-impact chronic LBP had more than doubled to 25.8% (8/111) since the 3-month follow-up (10.5%, 4/118). There were 6/111 (19.3%) new participants with chronic LBP at 6 months, 5 participants that had transitioned from a 3-month frequency of some days, and 1 participant that was missing 3 months outcomes that completed 6-month measures. There were 10 participants (12.5%) that had chronic LBP at 3 months but no longer were classified as chronic at 6 months. No participants that transitioned to chronic LBP answered “yes” to having a 30-day LBP-free period between baseline and 3 months or 3 months and 6 months. **Figure 1** illustrates the transition to and from chronic LBP over the 3 and 6 months. The proportion estimated from the NIH Task Force Minimum Data Set to have sought care for this episode of acute LBP was 26.1%. The treatments reported to be received did not influence the transition from acute to chronic LBP.

**Figure 1.**
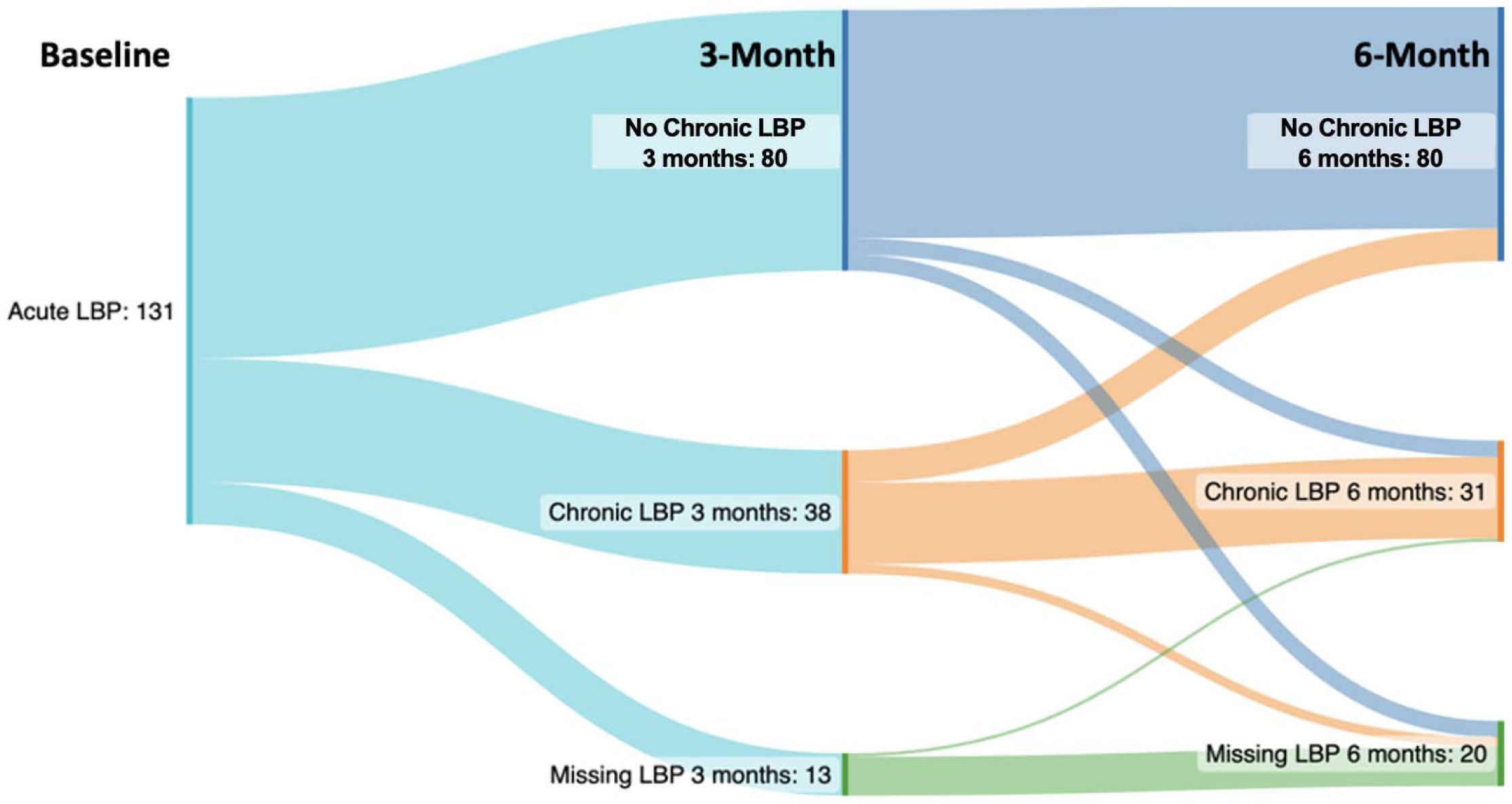
Sankey diagram of the transition to and from chronic LBP over 3 and 6 months.

**Figure 1.**
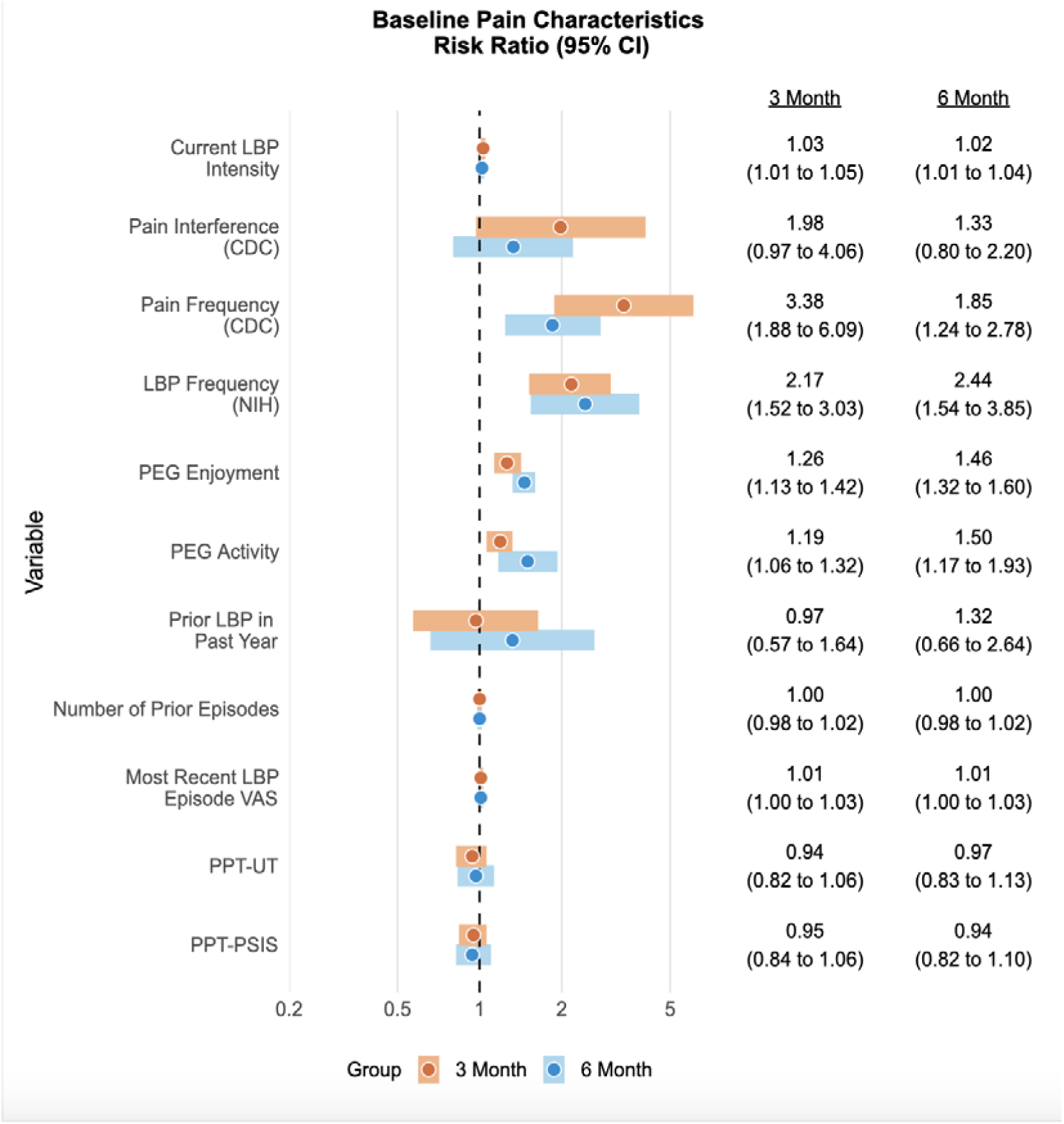
Risk Ratios for Baseline Pain Characteristics.

**Table 3** provides the differences in counts or means and effect sizes across baseline pain characteristics. Several pain characteristics reflected differences in participants who transitioned to chronic LBP at 3 and those continuing to have chronic LBP at 6 months. Current LBP intensity had a similar effect size at both 3 months (RR=1.03, 95% CI 1.01, 1.05) and 6 months (RR=1.02, 95% CI 1.01, 1.04). A similar relationship was found with the frequency of LBP over the past 4 weeks with those with half the days or less likely to have chronic LBP at 3 months (RR=0.46, 95% CI 0.33, 0.66) and 6 months (RR=0.41, 95% CI 0.26, 0.65). The PEG enjoyment scale was associated with those continuing to have chronic LBP at 6 months (RR=1.46, 95% CI 1.32, 1.60) and those transitioning at 3 months (RR=1.26, 95% CI 1.13, 1.42). Similarly, the PEG activity scale was found among those transitioning at 3 months (RR=1.26, 95% CI 1.13, 1.42) and those continuing to have chronic LBP at 6 months (RR=1.50, 95% CI 1.19, 1.88) **(Figure 1).**

Associations between age, sex, and overall race categories were similar among those transitioning to chronic LBP at 3 and those continuing to have chronic LBP at 6 months. When race was restricted to those self-reporting as Bl ck or White, Black participants were more likely to transition to chronic LBP at 3 months (RR=1.76, 95% CI 1.05, 2.95) and continue to have chronic LBP at 6 months (RR=2.03, 95% CI 1.11, 3.69). (**Figure 2**)

**Figure 2.**
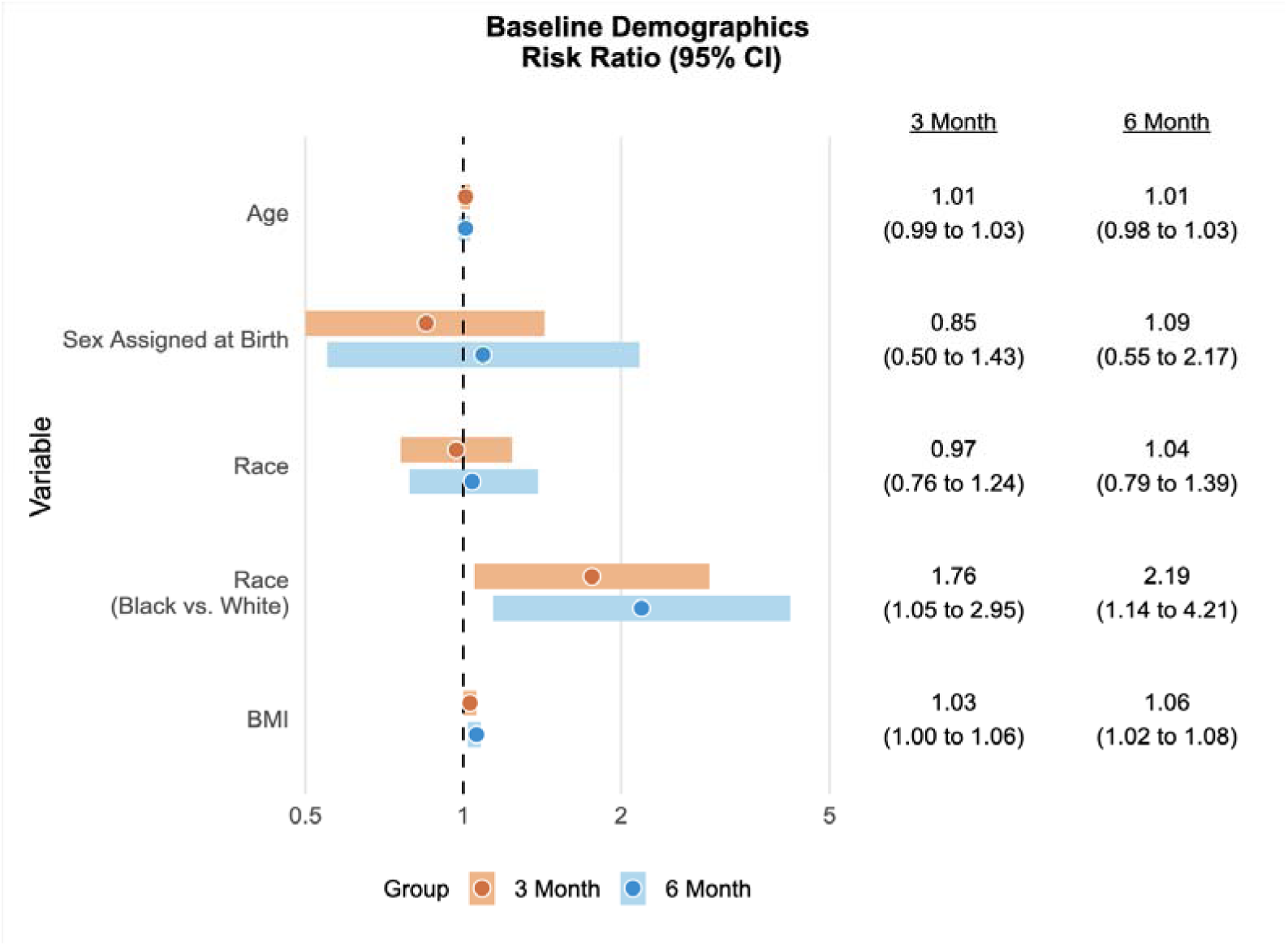
Baseline Demographic Risk ratios.

Those participants in the medium and high-risk categories of the STarT Back Screening Tool were more likely to transition to chronic LBP at 3 months (RR=1.73, 95% CI 1.28, 2.35) and continue to have chronic LBP at 6 months (RR=1.59, 95% CI 1.59, 2.36). In addition, participants with higher scores on the OSPRO-YF were more likely to transition to chronic LBP at 3 months (RR=1.11, 95% CI 1.03, 1.19) and continue to have chronic LBP at 6 mont s (RR=1.10, 95% CI 1.01, 1.19). (**Figure 3**.). Estimates for the unidimensional parent psychological measures from the OSPRO-YF can be found in the Supplementary Material.

**Figure 3.**
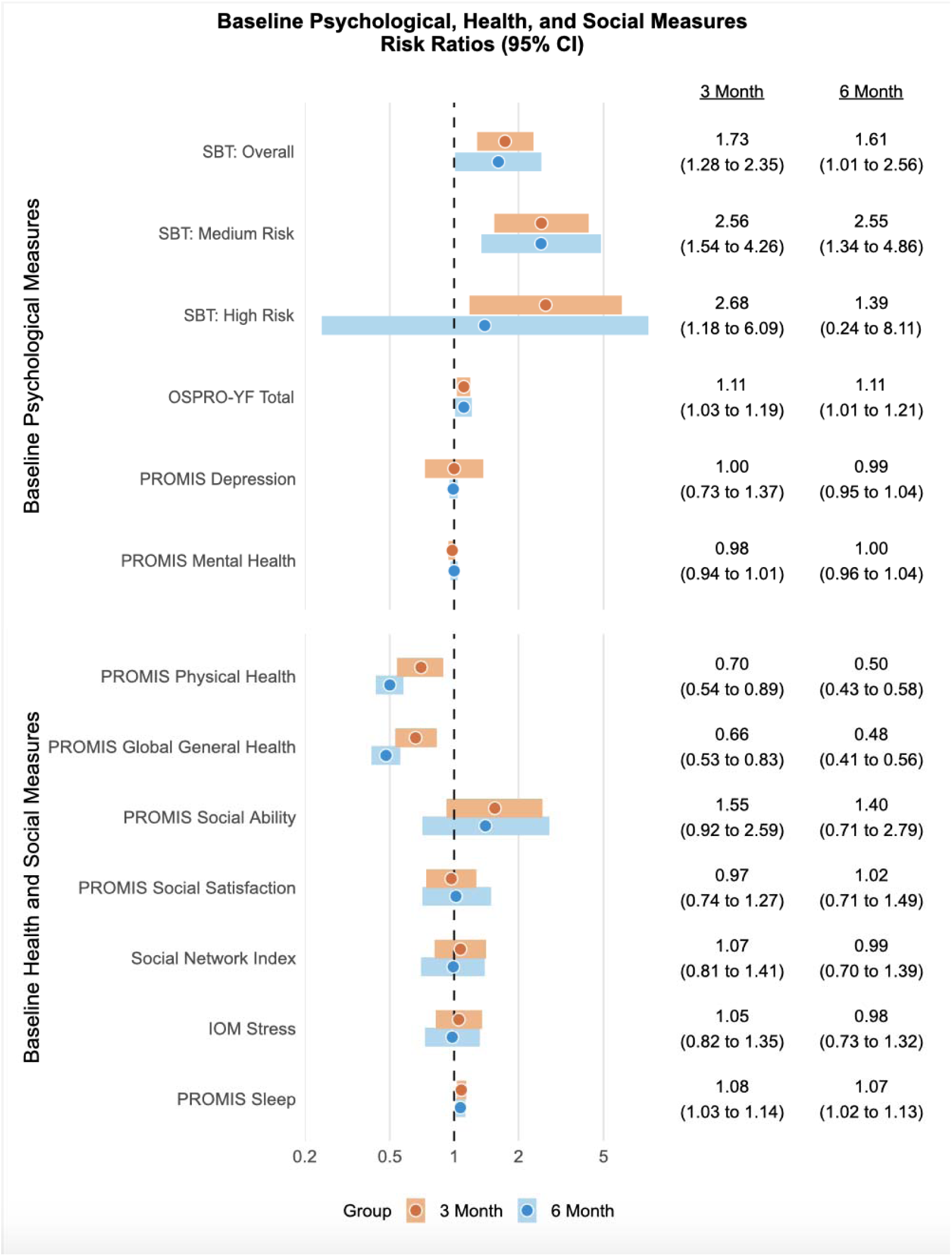
Risk Ratios for Baseline Psychological, Health and Social Measures.

**Figure 4.**
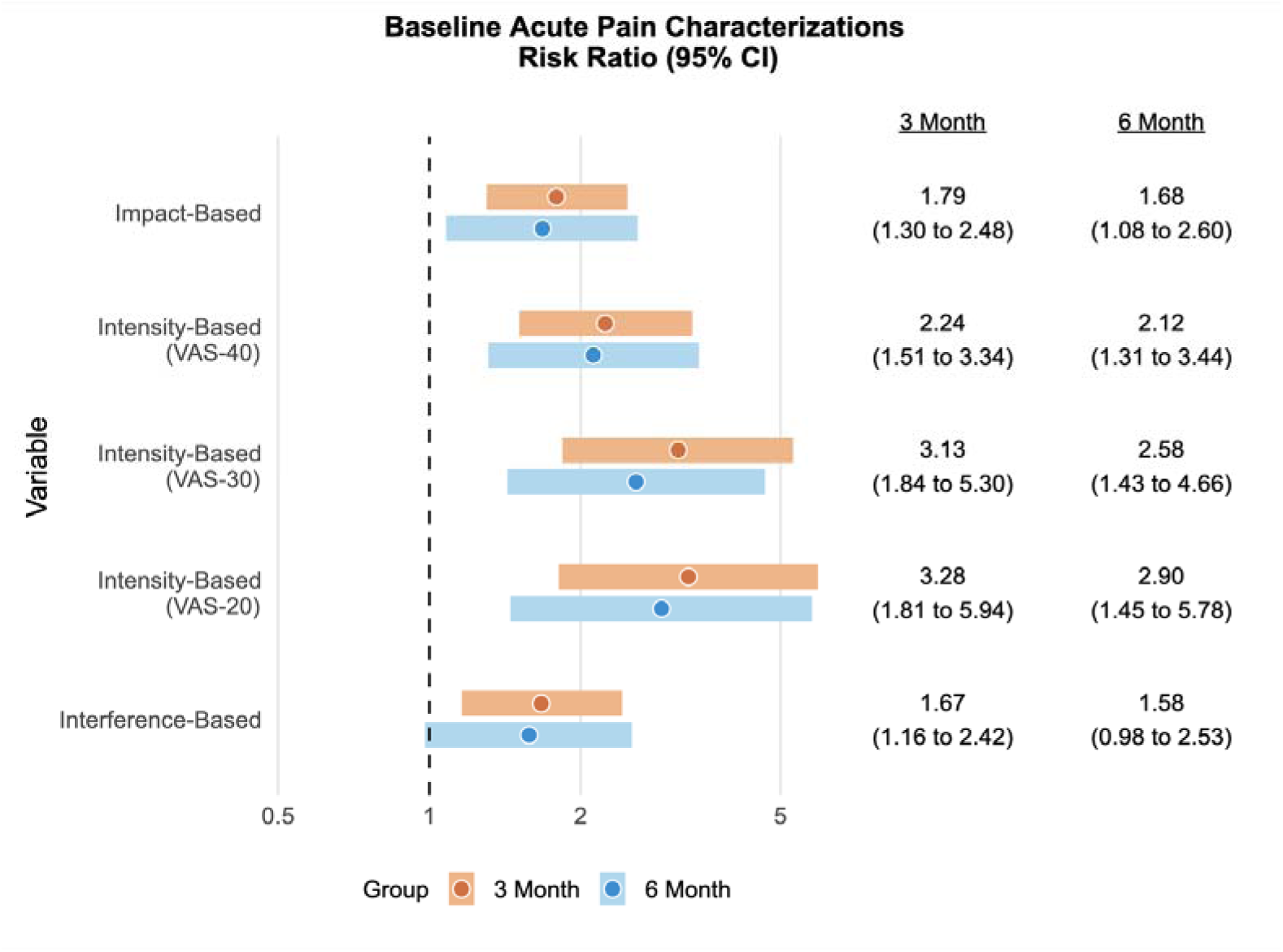
Risk Ratios for Baseline Acute Pain Characterizations.

Participants reporting better health on the PROMIS Global General Health scale were less likely to transition to chronic LBP at 3 months (RR=0.66, 95% CI 0.53, 0.83) and less likely to have continued chronic LBP at 6 months (RR=0.54, 95% CI 0.47, 0.62). Similarly, participants with better baseline physical health on the PROMIS Physical Health were less likely to transition to chronic LBP at 3 months (RR=0.70, 95% C 0.54, 0.89) and less likely to have chronic LBP at 6 months (RR=0.56, 95% CI 0.49, 0.65). Better baseline sleep scores on the PROMIS Sleep measure were less likely to transition to chronic LBP at 3 months (RR=1.08, 95% CI 1.03, 1.14) and less likely to have chronic LBP at 6 months (RR=1.07, 95% CI 1.02, 1.13). (**Figure 3**)

### Acute Pain definitions

Stronger associations were found with the acute LBP definitions that classified participants based on the intensity rather than frequency of LBP. Acute LBP classifications that used a 30/100 cut-off and 20/100 cut-off were strongly related to the transition to chronic LBP at 3 months (30/100, RR=3.13, 95% CI 1.84, 5.30; 20/100, RR=3.28, 95% CI 1.81, 5.94) and continuing chronic LBP at 6 months (30/100, RR=2.27, 95% CI 1.37, 3.77; 20/100, RR=2.61, 95% CI 1.43, 4.75).

## Discussion

We preliminary examined a diverse sample of community-based participants to determine the transition from acute LBP to chronic LBP at 3 and 6 months. In contrast to a recent study of community-based participants in Australia[15], we identified a relatively high incidence of chronic LBP at 3 months following a new acute episode of LBP. Interestingly, the transition rates reported in this cohort are consistent with a recent study of US patients seeking care [50] and potentially identify a “hidden” burden of chronic LBP among community-based participants who may not seek care in the first 6 months of a new LBP episode. This is important because previous studies have reported as many as 42% of community-based participants do not seek care for LBP[22]. In comparison, our estimates of participants utilizing treatments for LBP among our sample were much lower (26.1%). Although treatments during the acute stage would ideally prevent continued or progression of symptoms, the small proportion of participants we estimated to have sought care for acute LBP did not influence the transition to chronic LBP at 3 or prevent continued chronic LBP at 6 months. We also identified that a high proportion of those who transitioned (>80%) continue to have chronic LBP at 6 months, with a more than double proportion transitioning to high-impact chronic LBP at 6 months. This is in contrast to many of the prior studies that reported acute LBP as largely transient, with most experiencing resolution.[36; 44] While some studies have suggested different endpoints for estimating chronic LBP (3 months vs 6 months), our study demonstrates value in observing both time points when examining transitions. Some participants (19.6%) reported never or some days having LBP at 3 months and therefore did not meet the definition of chronic LBP, however then transitioned to having chronic LBP at 6 months. In addition, a relatively low proportion (12.5%) of participants transition from having chronic LBP at 3 months to not having chronic LBP at 6 months. These findings suggest that the impact of acute LBP is much greater than previously noted, at least among community-based participants, and is progressive, with more participants transitioning to worse chronic LBP and only a limited proportion of participants improving over time.

Sociodemographic characteristics and social factors are known to influence health and quality-of-life outcomes.[1; 2] Our study identified that those participants who self-identify as Black or African American race are more likely to transition to chronic LBP at 3 months and continue to have chronic symptoms at 6 months. While racial disparities are known to exist in the prevalence, symptoms, and treatment of chronic LBP, these findings are unique to community-based studies but are supported by race differences in the cross-sectional analysis of participants with chronic LBP.[11; 12] Interestingly, we did not find any differences in sex or age for the transition to chronic LBP at 3 months or continued chronic LBP at 6 months despite other studies that have reported a higher proportion of women who report chronic LBP. One reason for this may be the close to equal proportion of women and men in the study, which typically would reflect a higher proportion of women in care-seeking populations. Women are more likely to seek health care for LBP and do so more frequently compared to men[20; 33], which could be indicative of higher levels of health-seeking behaviors among women. Since we did not rely on care-seeking populations and noticed similar transition rates for men and women, our results present a unique perspective on the association between sex and gender and the transition from acute to chronic LBP.

Furthermore, general health (i.e., physical and mental health) and social well-being are determined by a number of factors both within and outside of the healthcare system.[38] Better general health, physical health, and sleep quality were protective factors during the acute episode of LBP associated with a decreased risk of transitioning to chronic LBP. We viewed these protective findings as novel since identifying potential preventive factors has not been the focus of prior studies on the transition to chronic LBP. Data from acute pain stages presents a unique opportunity to identify early indicators of chronic pain vulnerability and inform targeted interventions. Identifying protective elements against the progression of acute LBP to the chronic stage represents a paradigm shift in pain management, emphasizing the importance of a comprehensive, multidimensional approach. Integrating interventions promoting healthy lifestyles to improve general health, social well-being, and sleep hygiene may mitigate the risk of chronic pain development. By elucidating the protective mechanisms that mitigate the transition from acute to chronic LBP, researchers and clinicians can proactively enhance early detection to optimize treatment strategies.

Other baseline social measures, including social isolation, social ability, and social satisfaction, were not associated with the transition from acute to chronic LBP. However, a recent study examined the correlation between overall health (PROMIS Global Health) and social deprivation among orthopaedic surgical patients[5]. In unadjusted models, there was a poor correlation between overall health and social deprivation, but when adjusting for sociodemographic factors, area deprivation indices were associated with both physical and mental health scores. These findings may suggest that the initial acute pain episodes may not be enough for these social factors to impact persistence of pain. Instead, it might be that social factors and pain interact over time, which is why they are seen as stronger contributors for those who have chronic pain because they have experienced pain for longer. This suggests the need for further analyses on the effects of our social factors over time, as well as any associations between subgroups like sociodemographic characteristics or acute pain categorizations, which may help explain the complexities of the relationship between social factors and the development of chronic LBP as it did in the orthopedic surgical patients.

Psychological distress is a well-established risk factor for chronic LBP. Similar to prior studies examining chronic LBP and the transition from acute to chronic LBP among care-seeking patients, we identified both the STarT Back Screening Tool (SBT) and the OSPRO-YF associated with the transition to chronic LBP and the continuation of LBP. Although both measure aspects of psychological distress, the two measures may capture different aspects of distress, and there may be added value to using both to identify risk for transition.[40] Further analyses are warranted to determine the benefits and combined predictive capabilities of these measures in the community setting. Within the OSPRO-YF domains, pain resiliency was not associated with the transition to chronic pain. This is similar to previous findings, and may be due to the duration and timing of this study.[21] Resilience may be an important factor in promoting adaptation to chronic pain, but play a limited role in preventing or limiting the transition to chronic pain. Interestingly, PROMIS Depression did not demonstrate significant associations with the transition to chronic pain, however there is some evidence that PROMIS Depression may not be adequately capturing depression symptoms in orthopedic populations[6] and the scores were generally within the normal ranges. This contrasts the findings on both the SBT and OSPRO-YF, which has greater specificity for pain, which the PROMIS Depression may not capture. These findings support the need for appropriate and thorough screening in this population that may be pain-specific, as participants in the acute phase may not have had exposure to the painful stimulus persistent enough to be linked to negative mood.

We are unaware of prior studies that have examined the different definitions of acute LBP based on the frequency and/or intensity of baseline symptoms and the transition to chronic LBP at 3 months and continued chronic LBP at 6 months. Prior studies discussing pain distributions have focused exclusively on categorizing chronic pain and examining the natural history of chronic pain [17; 54], but only examined these categorizations within the chronic state. These categorizations have yet to be studied from an acute to a chronic stage, likely because an established definition of acute LBP categorizations does not exist. A recent review has suggested utilizing both the frequency and intensity of LBP as a framework for examining both acute and chronic pain, as well as the transition between states of pain.[18] Indeed, our ongoing work has identified that different definitions of acute LBP by baseline frequency, impact, intensity, and interference may influence relationships with psychosocial factors. [52] We examined these definitions in relationship with those who transition to chronic LBP and those with continued chronic LBP symptoms at 6 months. The use of both intensity of LBP and frequency of LBP, specifically at the 30/100 or 40/100 cut point of intensity, appear to be meaningfully different in their association with chronic LBP at 3 months. These findings were also consistent with both the 3-month transition to chronic LBP and as an indication for those participants who continued to have chronic symptoms at 6 months. These findings support the need for standardized definitions of acute LBP that appear to have a substantial influence on identifying those individuals who go on to transition to chronic LBP and continue to have chronic LBP symptoms at 6 months.

This study has several strengths, including comprehensive psychological and social measurements and a community-based cohort sample. However, there are limitations to this study. First, the primary purpose of this study was to collect preliminary data to assess the feasibility of recruiting acute LBP participants within the community and follow these participants longitudinally. Our sample size limited the ability to examine all categories of chronic LBP (i.e., high-impact, bothersome, and mild) and the potential reasons for those participants who transitioned from mild or bothersome chronic LBP to high-impact LBP between 3 and 6 months. Future studies with larger sample sizes should examine this important transition to identify potential subgroups for treatments. Our sample size was limited, impacting the precision of our estimates. The small sample size may impact the generalizability of our findings. Second, although we excluded participants who did not have 30 consecutive days without LBP before the onset of their current LBP episode to restrict our cohort to potential recurrent acute LBP, however, some participants reported other LBP episodes within the past year. Some participants may meet the consensus definition for recurrent LBP, depending on the context surrounding these previous LBP episodes (which occurred and resolved ≥30 days before the current LBP episode).[49] However, our fielded questions regarding pain-free periods between 3- and 6-months support that those individuals having pain-free periods are unlikely to transition to chronic LBP. Despite these limitations, our study comprehensively characterizes acute LBP within the community and provides early estimates of the transition and continuation of chronic LBP.

## Conclusion

This preliminary study identified that the transition to chronic LBP for those in the community was comparable to that of care-seeking populations in the US. The transition to high-impact chronic LBP from 3 to 6 months more than doubled, and relatively few participants in this cohort reported improved symptoms over the 6 month follow-up. The categorization of acute LBP, general and physical health, psychological factors, and self-identified race at baseline have important implications for studies seeking to determine the transition from acute to chronic LBP. Future studies are needed to understand better the reasons for the differences in race in the transition to chronic LBP. Studies with larger sample sizes are needed to improve the generalizability of these findings.

## Data Availability

All data produced in the present study are available upon reasonable request to the authors

