## Supplementary Table for "Transition from Acute to Chronic Low Back Pain in a Community-Based Cohort"

**Supplementary Table 1. Differences in baseline pain characteristics by transition at 3 and continuation of LBP at 6 months.**

|  | **Transition to Chronic LBP at 3-months** | | | **Continued Chronic LBP at 6-months** | | |
| --- | --- | --- | --- | --- | --- | --- |
|  | Yes (n=38) | No (n=80) | p-value | Yes (n=26) | No (n=80) | p-value |
| Current LBP Intensity | 48.8 (17.4) | 35.7 (21.8) | 0.002 | 48.2 (19.4) | 35.5 (21.0) | 0.008 |
| Pain Frequency (CDC) |  |  | <0.0001 |  |  | 0.003 |
| Some Days | 7 (18.4%) | 50 (62.5%) |  | 5  (19.2%) | 46 (57.5%) |  |
| Most Days | 20 (52.6%) | 22 (27.5%) |  | 14  (53.9%) | 24 (30.0%) |  |
| Every Day | 11 (29.0%) | 8 (10.0%) |  | 7  (26.9%) | 10 (12.5%) |  |
| Pain Interference (CDC) |  |  | 0.04 |  |  | 0.19 |
| Never | 3  (7.9%) | 20 (25.0%) |  | 2  (7.7%) | 20 (25.0%) |  |
| Some Days | 30 (79.0%) | 55 (68.8%) |  | 21 (80.8%) | 53 (66.3%) |  |
| Most Days | 5 (13.2%) | 3 (3.8%) |  | 3  (11.5%) | 5  (6.3%) |  |
| Every Days | 0  (0.0%) | 2  (2.5%) |  | 0  (0.0%) | 2  (2.5%) |  |
| LBP Frequency (NIH) |  |  | <0.0001 |  |  | >0.0001 |
| Every day or nearly every day past 4 weeks | 17 (44.7%) | 11 (13.8%) |  | 11 (42.3%) | 12 (15.0%) |  |
| At least half the days past 4 weeks | 17 (44.7%) | 33 (41.3%) |  | 14 (53.9%) | 32 (40.0%) |  |
| Less than half the days in past 4 weeks | 4 (10.5%) | 36 (45.0%) |  | 1  (3.9%) | 36 (45.0%) |  |
| PEG Enjoyment, mean (SD) | 4.9 (1.98) | 3.2 (1.98) | >0.0001 | 4.8  (2.1) | 3.1  (1.9) | 0.0004 |
| PEG Activity, mean (SD) | 4.8 (1.8) | 3.3 (2.2) | 0.0005 | 4.7  (1.8) | 3.2  (2.0) | 0.0007 |
| Prior LBP in past year, n (%) |  |  | 0.91 |  |  | 0.42 |
| No | 18 (47.4%) | 37 (46.3%) |  | 10 (38.5%) | 38 (47.5%) |  |
| Yes | 20 (52.6%) | 43 (53.8%) |  | 16  (61.5%) | 42 (52.5%) |  |
| Number of Prior Episodes (n=69) |  |  | 0.36 |  |  | 0.27 |
| 1 | 1 (5.0%) | 5 (11.6%) |  | 1  (6.3%) | 5 (11.9%) |  |
| 2 | 5 (25.0%) | 11 (25.6%) |  | 4 (25.0%) | 10 (23.8%) |  |
| 3 | 7 (35.0%) | 7 (16.3%) |  | 4 (25.0%) | 9 (21.4%) |  |
| 4+ | 7 (35.0%) | 20 (46.5%) |  | 8  (50.0%) | 18 (42.9%) |  |
| Most recent LBP episode VAS (n=69) | 56.2 (17.2) | 49.7 (22.7) | 0.26 | 58.7 (19.8) | 49.5 (22.1) | 0.15 |
| PPT-UT | 4.5 (2.3) | 5.0 (2.2) | 0.33 | 4.7  (2.4) | 4.9  (2.2) | 0.71 |
| PPT-PSIS | 4.7 (2.4) | 5.2 (2.4) | 0.36 | 4.7  (2.3) | 5.1  (2.4) | 0.44 |

LBP=low back pain, CDC=Centers for Disease Control, NIH=National Institutes of Health, SD=standard deviation, PEG=pain, enjoyment, general activity scale,

**Supplementary Table 2. Associations between baseline demographics, acute low back pain definitions, and the transition to chronic low back pain at 3 and 6 months**.

|  | **Transition to Chronic LBP at 3-months** | | | **Continued Chronic LBP at 6-months** | | |
| --- | --- | --- | --- | --- | --- | --- |
| **Demographic Characteristics** | Yes (n=38) | No (n=80) | p-value | Yes (n=26) | No (n=80) | p-value |
| **Age** | 59.1 (1.9) | 57.1 (1.5) | 0.46 | 59.3  (11.2) | 58.1  (13.5) | 0.67 |
| **Sex at Birth** |  |  | 0.38 |  |  | 0.80 |
| Female | 21(56.3%) | 21  (30%) |  | 16  (61.5%) | 47 (71.2%) |  |
| Male | 17(44.7%) | 49  (70%) |  | 10 (38.5%) | 33 (73.3%) |  |
| **Race** |  |  | 0.21 |  |  | 0.15 |
| White | 23 (28.8%) | 57 (71.3%) |  | 19 (23.8%) | 61 (76.3%) |  |
| Black or African American | 13 (50%) | 13  (50%) |  | 10 (47.6%) | 11 (52.4%) |  |
| Multiple Races | 1  (20%) | 4  (80.0%) |  | 4 (100.0%) | 0.0 (0.0%) |  |
| Asian | 0  (0%) | 2  (100.0%) |  | 1 (50.0%) | 1  (50.0%) |  |
| Unknown | 1 (20.0%) | 4  (80.0%) |  | 1 (25.0%) | 3  (75.0%) |  |
| White | 23 (28.4%) | 58 (71.6%) | 0.04 | 16 (20.5%) | 62 (79.5%) | 0.03 |
| Black or African American | 13 (50%) | 13  (50%) |  | 9 (45.0%) | 11 (55.0%) |  |
| **BMI**  Mean (SD) | 32.0 (1.3) | 29.5 (0.71) | 0.07 | 31.8 (8.3) | 29.1 (5.5) | 0.06 |
| **Acute LBP Categorizations** |  |  |  |  |  |  |
| **Frequency-Based** |  |  | 0.009 |  |  | 0.007 |
| Mild | 16  (42.1%) | 57  (71.3%) |  | 11  (42.3%) | 58  (72.5%) |  |
| Bothersome | 18  (47.4%) | 18  (22.5%) |  | 13  (50.0%) | 15  (18.8%) |  |
| High | 4  (10.5%) | 5  (6.3%) |  | 2  (7.7%) | 7  (8.8%) |  |
| **Impact-Based** |  |  | <0.0001 |  |  | <0.0001 |
| Low | 8 (21.1%) | 55 (68.8%) |  | 6  (23.1%) | 51 (63.8%) |  |
| Medium | 26 (68.4%) | 20 (25.0%) |  | 18 (69.2%) | 22 (27.5%) |  |
| High | 4 (10.5%) | 5  (6.3%) |  | 2  (7.7%) | 7  (8.8%) |  |
| **Intensity-Based 40** |  |  | <0.0001 |  |  | 0.005 |
| Weak | 3  (7.9%) | 30 (37.5%) |  | 3  (9.4%) | 29 (36.3%) |  |
| Moderate | 13 (34.2%) | 33 (41.3%) |  | 8 (30.1%) | 31 (38.8%) |  |
| Strong | 22 (57.9%) | 17 (21.3%) |  | 15 (57.7%) | 20 (25.0%) |  |
| **Intensity-Based 30** |  |  | <0.0001 |  |  | 0.001 |
| Weak | 3  (7.9%) | 23 (28.8%) |  | 3  (11.5%) | 23 (28.8%) |  |
| Moderate | 5 (13.2%) | 36 (45.0%) |  | 3  (11.5%) | 31 (38.8%) |  |
| Strong | 30 (79.0%) | 21 (26.3%) |  | 20 (76.9%) | 26 (32.5%) |  |
| **Intensity-Based 20** |  |  | <0.0001 |  |  | 0.002 |
| Weak | 1  (2.6%) | 17 (21.3%) |  | 1  (3.9%) | 16 (20.0%) |  |
| Moderate | 7 (18.4%) | 37 (46.3%) |  | 5  (19.2%) | 34 (42.5%) |  |
| Strong | 30 (79.0%) | 26 (32.5%) |  | 20 (76.9%) | 30 (37.5%) |  |
| **Interference-Based** |  |  | 0.01 |  |  | 0.02 |
| Minimal | 10 (26.3%) | 44 (55.0%) |  | 7 (26.9%) | 45 (56.3%) |  |
| Intermediate | 24 (63.2%) | 32 (40.0%) |  | 17 (65.4%) | 29 (36.3%) |  |
| Frequent | 4 (10.5%) | 4  (5.0%) |  | 2  (7.7%) | 6  (7.5%) |  |

BMI=body mass index, SD=standard deviation, LBP=low back pain,

**Supplementary Table 3. Baseline Psychological Measures and the transition to chronic LBP at 3 months and continuation of LBP at 6-months.**

|  | **Transition to Chronic LBP at 3-months** | | | **Continued Chronic LBP at 6-months** | | |
| --- | --- | --- | --- | --- | --- | --- |
|  | Yes (n=38) | No (n=80) | p-value | Yes (n=26) | No (n=80) | p-value |
| Start Back Tool |  |  | 0.001  0.001 (Exact) |  |  | 0.026  0.020 (Exact) |
| Low Risk | 19 (50.0%) | 66 (82.5%) |  | 14 (53.9%) | 64 (80.0%) |  |
| Medium Risk | 16 (42.1%) | 12 (15.0%) |  | 11 (42.3%) | 13 (16.3%) |  |
| High Risk | 3 (7.9%) | 2 (2.5%) |  | 1  (3.9%) | 3 (3.8%) |  |
| OSPRO-YF-Total | 5.45 (3.16) | 3.5 (3.2) | 0.003 | 5.42 (3.14) | 3.67 (3.34) | 0.02 |
| PROMIS Depression | 46.5 (6.6) | 47.2 (8.3) | 0.67 | 46.2 (6.0) | 46.6 (7.4) | 0.80 |
| PROMIS Mental Health | 49.4 (8.3) | 52.3 (8.1) | 0.15 | 52.2 (8.1) | 51.9 (8.4) | 0.88 |

OPSRO-YF=Optimal Screening for Prediction of Referral and Outcome Yellow Flag tool

**Supplementary Table 4. Baseline health and social health measures and the transition to chronic low back pain at 3 months and continuation of LBP at 6 months.**

|  | **Transition to Chronic LBP at 3-months** | | | **Continued Chronic LBP at 6-months** | | |
| --- | --- | --- | --- | --- | --- | --- |
| Chronic LBP | Yes (n=38) | No  (n=80) | p-value | Yes (n=26) | No (n=80) | p-value |
| **PROMIS Global General Health** |  |  | 0.017 |  |  | 0.014 |
| Poor | 2  (5.3%) | 1  (1.3%) |  | 2  (7.7%) | 0  (0%) |  |
| Fair | 6 (15.8%) | 5  (6.3%) |  | 4 (15.4%) | 6  (7.5%) |  |
| Good | 20 (52.6%) | 30 (37.5%) |  | 13 (50.0%) | 30  (37.5) |  |
| Very Good | 10 (26.3%) | 34  (42.5) |  | 7 (16.7%) | 35 (43.8%) |  |
| Excellent | 0  (0%) | 10 (12.5%) |  | 0  (0.0%) | 9  (11.3) |  |
| **Social Ability** |  |  | 0.11 |  |  | 0.26 |
| Excellent or Very Good | 23 (60.5%) | 60 (75.0%) |  | 17 (65.4%) | 60  (75%) |  |
| Poor, Fair or Good | 15 (39.5%) | 20 (25.0%) |  | 9 (34.6%) | 20 (25.0%) |  |
| **Social Satisfaction** |  |  |  |  |  |  |
| Poor | 0  (0.0%) | 1  (1.3%) |  | 0  (0.0%) | 1  (1.3%) |  |
| Fair | 6  (16.2%) | 12  (15.4%) |  | 2  (8.0%) | 9  (11.5%) |  |
| Good | 9  (24.3%) | 3  (16.7%) |  | 8  (32.0%) | 14  (18.0%) |  |
| Very Good | 15  (40.5%) | 38  (48.7%) |  | 9  (36.0%) | 40  (51.3%) |  |
| Excellent | 7  (18.9%) | 14  (18.0%) |  | 6  (24.0%) | 14  (18.0%) |  |
| **PROMIS Physical Health** |  |  | 0.12 |  |  | 0.005 |
| Poor | 2  (5.4%) | 1  (1.3%) |  | 2  (7.7%) | 0  (0.0%) |  |
| Fair | 5  (13.5%) | 6  (7.5%) |  | 2  (7.7%) | 7  (8.8%) |  |
| Good | 18  (48.7%) | 28  (35.0%) |  | 16  (61.5%) | 27  (33.8%) |  |
| Very Good | 11  (29.7%) | 37  (46.3%) |  | 5  (19.2%) | 39  (48.8%) |  |
| Excellent | 1  (2.7%) | 8  (10.0%) |  | 1  (3.9%) | 7  (8.8%) |  |
| **Stress** |  |  | 0.60 |  |  | 0.97 |
| Not at all | 12 (32.4%) | 22 (27.5%) |  | 8 (30.7%) | 24 (34.0%) |  |
| A little bit | 13 (35.1%) | 38 (47.5%) |  | 11 (42.3%) | 36 (45.0%) |  |
| Somewhat | 6 (16.2%) | 12 (15.0%) |  | 4 (15.4%) | 11 (13.8%) |  |
| Quite a bit | 5 (13.5%) | 5  (6.3%) |  | 3  (11.5%) | 7  (8.8%) |  |
| Very Much | 1  (2.7%) | 3  (3.8%) |  | 0  (0.0%) | 2  (2.5%) |  |
| **Isolation** |  |  | 0.91 |  |  | 0.93 |
| Most Isolated | 6 (16.2%) | 13 (16.3%) |  | 7 (22.6%) | 12 (15.0%) |  |
| Very Isolated | 6 (16.2%) | 17 (21.3%) |  | 5 (16.1%) | 17 (21.3%) |  |
| Somewhat Isolated | 16 (43.2%) | 34 (42.5%) |  | 12 (38.7%) | 34 (42.5%) |  |
| Not Isolated | 9 (24.3%) | 16 (20.0%) |  | 7 (22.6%) | 17 (21.3%) |  |
| **PROMIS Sleep** | 53.3  (8.7) | 48.0  (8.1) | 0.001 | 53.1  (8.9) | 48.1  (8.3) | 0.006 |

LBP=low back pain

**Supplementary Table 5. Transition to chronic low back pain by acute low back pain definitions at 3 months and continued LBP at 6 months.**

|  | **Transition to Chronic LBP at 3-months** | | | **Continued Chronic LBP at 6-months** | | |
| --- | --- | --- | --- | --- | --- | --- |
| Chronic LBP | Yes (n=38) | No  (n=80) | p-value | Yes (n=26) | No (n=80) | p-value |
| **Acute LBP Categorizations** |  |  |  |  |  |  |
| **Impact-Based** |  |  | <0.0001 |  |  | <0.0001 |
| Low | 8 (21.1%) | 55 (68.8%) |  | 6  (23.1%) | 51 (63.8%) |  |
| Medium | 26 (68.4%) | 20 (25.0%) |  | 18 (69.2%) | 22 (27.5%) |  |
| High | 4 (10.5%) | 5  (6.3%) |  | 2  (7.7%) | 7  (8.8%) |  |
| **Intensity-Based 40** |  |  | <0.0001 |  |  | 0.005 |
| Weak | 3  (7.9%) | 30 (37.5%) |  | 3  (9.4%) | 29 (36.3%) |  |
| Moderate | 13 (34.2%) | 33 (41.3%) |  | 8 (30.1%) | 31 (38.8%) |  |
| Strong | 22 (57.9%) | 17 (21.3%) |  | 15 (57.7%) | 20 (25.0%) |  |
| **Intensity-Based 30** |  |  | <0.0001 |  |  | 0.001 |
| Weak | 3  (7.9%) | 23 (28.8%) |  | 3  (11.5%) | 23 (28.8%) |  |
| Moderate | 5 (13.2%) | 36 (45.0%) |  | 3  (11.5%) | 31 (38.8%) |  |
| Strong | 30 (79.0%) | 21 (26.3%) |  | 20 (76.9%) | 26 (32.5%) |  |
| **Intensity-Based 20** |  |  | <0.0001 |  |  | 0.002 |
| Weak | 1  (2.6%) | 17 (21.3%) |  | 1  (3.9%) | 16 (20.0%) |  |
| Moderate | 7 (18.4%) | 37 (46.3%) |  | 5  (19.2%) | 34 (42.5%) |  |
| Strong | 30 (79.0%) | 26 (32.5%) |  | 20 (76.9%) | 30 (37.5%) |  |
| **Interference-Based** |  |  | 0.01 |  |  | 0.02 |
| Minimal | 10 (26.3%) | 44 (55.0%) |  | 7 (26.9%) | 45 (56.3%) |  |
| Intermediate | 24 (63.2%) | 32 (40.0%) |  | 17 (65.4%) | 29 (36.3%) |  |
| Frequent | 4 (10.5%) | 4  (5.0%) |  | 2  (7.7%) | 6  (7.5%) |  |

LBP=low back pain

**Supplementary Table 6. Estimates of psychological distress from the OSPRO-YF tool for transition to chronic LBP at 3 months and the continuation of chronic LBP at 6 months.**

|  | Transition to chronic LBP at 3-months | |  | Continuation of chronic LBP at 6-months | |  |
| --- | --- | --- | --- | --- | --- | --- |
|  | Yes (n=38) | No (n=80) | p-value | Yes (n=31) | No (n=80) | p-value |
| **PHQ9** | 6.1 (4.1) | 4.2 (3.5) | 0.01 | 5.31 (3.3) | 4.32 (3.8) | 0.21 |
| **STAI** | 37.3 (6.8) | 34.4 (6.3) | 0.03 | 36.0 (6.0) | 34.6 (6.6) | 0.31 |
| **STAXI** | 15.2 (2.5) | 14.3 (2.4) | 0.06 | 14.6 (2.2) | 14.4 (2.5) | 0.61 |
| **FABQ-PA** | 15.0 (3.5) | 12.0 (4.1) | 0.0001 | 15.1 (3.7) | 12.4 (4.2) | 0.002 |
| **FABQ-W** | 11.8 (7.5) | 8.9 (7.0) | 0.046 | 12.2 (8.4) | 8.8 (6.3) | 0.025 |
| **PCS** | 15.1 (7.7) | 10.9 (7.5) | 0.0059 | 14.8 (7.8) | 11.0 (7.7) | 0.02 |
| **TSK-11** | 23.1 (4.1) | 20.8 (4.1) | 0.0034 | 23.4 (4.4) | 20.9 (3.9) | 0.0041 |
| **PASS20** | 28.7 (11.0) | 22.3 (11.3) | 0.0044 | 28.6 (11.2) | 22.4 (11.5) | 0.012 |
| **PSEQ** | 36.2 (7.2) | 39.2 (10.2) | 0.11 | 36.7 (6.9) | 38.8 (10.5) | 0.32 |
| **SER** | 88.4 (15.2) | 91.5 (20.4) | 0.41 | 90.2 (14.9) | 90.9 (20.4) | 0.86 |
| **CPAQ** | 64.2 (11.4) | 68.5 (14.4) | 0.11 | 64.9 (10.7) | 68.2 (14.9) | 0.26 |

LBP=low back pain, PHQ9=patient health questionnaire 9, STAI=State Trait Anxiety Inventory, STAXI=State Trait Anger Expression Inventory, FABQ-PA=Fear Avoidance Behavior Questionnaire Physical Activity, FABQ-W=Fear Avoidance Behavior Questionnaire Work, PCS=Pain Catastrophizing Scale, TSK-11=Tampa Scale, PASS20=Pain Anxiety Symptoms Scale, PSEQ=Pain Self Efficacy Questionnaire, SER=Self-Efficacy for Rehabilitation, CPAQ=Chronic Pain Acceptance Questionnaire.
